# Antibody responses to BNT162b2 mRNA COVID-19 vaccine in 2,015 healthcare workers in a single tertiary referral hospital in Japan

**DOI:** 10.1101/2021.06.01.21258188

**Authors:** Takahiro Kageyama, Kei Ikeda, Shigeru Tanaka, Toshibumi Taniguchi, Hidetoshi Igari, Yoshihiro Onouchi, Atsushi Kaneda, Kazuyuki Matsushita, Hideki Hanaoka, Taka-Aki Nakada, Seiji Ohtori, Ichiro Yoshino, Hisahiro Matsubara, Toshinori Nakayama, Koutaro Yokote, Hiroshi Nakajima

## Abstract

We measured antibody responses in 2,015 healthcare workers who were receiving 2 doses of BNT162b2 mRNA vaccine against SARS-CoV-2. The vast majority (99.9%) had either seroconversion or a substantial increase in antibody titer. A multivariate linear regression model identified predictive factors for antibody responses which may have clinical implications.

## Introduction

BNT162b2 mRNA vaccine against COVID-19 has shown promising efficacy both in the clinical trial [1] and in nationwide mass vaccination settings [2]. The vaccine has also shown short-term efficacy in a large-scale prospective cohort study targeting healthcare workers, a population that should be prioritized to be vaccinated [3]; however, there remains uncertainty as regards the long-term protection, the prediction of effectiveness, and the optimal usage of BNT162b2 mRNA vaccine.

As the humoral responses have been shown to play essential roles in the protection against and the survival from SARS-CoV-2 infection [4, 5], the antibody status after vaccination can provide vital information to predict long-term effectiveness and to optimize the vaccination strategy. However, antibody responses after vaccination in a real-world setting have only been shown in several small-scale studies [6, 7]. Here, we report the antibody responses and their predictive factors in 2,015 healthcare workers who received the BNT162b2 mRNA COVID-19 vaccine.

## Methods

We recruited healthcare workers in Chiba University Hospital who were receiving the BNT162b2 mRNA COVID-19 vaccine (Pfizer, Inc., and BioNTech) in our hospital vaccination program. All participants gave written informed consent before undergoing any study procedures.

Background information was collected by a web-based questionnaire. Blood samples were obtained 0-2 weeks before the 1^st^ dose and 2-5 weeks after the 2^nd^ dose of vaccination. Antibody responses were analyzed using Elecsys® Anti-SARS-CoV-2S on Cobas 8000 e801 module (Roche Diagnostics, Rotkreuz, Switzerland). This system allows for the quantitative detection of antibodies, predominantly IgG, aiming at the SARS-CoV-2 spike protein receptor-binding domain. Values ≥0.8 U/mL were considered positive. Samples with a titer >250 U/mL were diluted to determine the value.

Antibody titers were converted to log 2 values for presentation and statistical analysis. Univariate linear regression was performed using a forced entry method, whereas a multivariate linear regression model was established using a stepwise method. Statistical analyses were performed using SPSS version 23.0 (IBM, Armonk, NY). A two-sided p-value <0.05 was considered statistically significant.

The study procedures for sample collection and those for analyses were approved by Chiba University Ethics Committee on February 24^th^, 2021 (No. HS202101-03) and April 21^st^, 2021 (No. HS202104-01), respectively.

## Results

Out of 2,838 employees in Chiba University Hospital, 2,549 received at least one dose of BNT162b2 mRNA COVID-19 vaccine (30 µg) from March 3^rd^ to April 9^th^, 2021, and 2,015 individuals were enrolled in this study.

Median age was 37 year-old (interquartile range [IQR] 29 – 47) and 1,296 (64.3%) were women. All participants but 11 were Japanese (99.5%). Nurses (n=672) and doctors (n=591) comprised about two-thirds of the study population. Ten participants (0.5%) had a history of COVID-19 confirmed by polymerase chain reaction testing. Detailed background information is provided in Supplementary Table S1.

Before vaccination, the serum anti-SARS-CoV-2S antibody was detected (≥0.4 U/mL) only in 21 subjects (1.1%) with a median titer of 35.9 U/mL (IQR 7.8 – 65.7). Eighteen subjects (0.9%) had a positive titer (≥0.8 U/mL) and 8 out of these 18 subjects (44.4%) had a history of COVID-19.

After vaccination, the serum anti-SARS-CoV-2S antibody was detected in all 1,774 participants who received the 2^nd^ dose with a median titer of 2060.0 U/mL [1250.0-2650.0] (Figure 1A). Only one subject, who had received aggressive immunosuppressive treatment for a severe autoimmune condition, had a negative titer (0.7 U/mL). The distribution of post-vaccination antibody titers according to age and sex is shown in Figure 1B. In those who were seropositive before vaccination, the antibody titers substantially increased with a median fold change of 412.4 (IQR 309.2 – 760.5) following the 2^nd^ dose.

**Figure 1.**
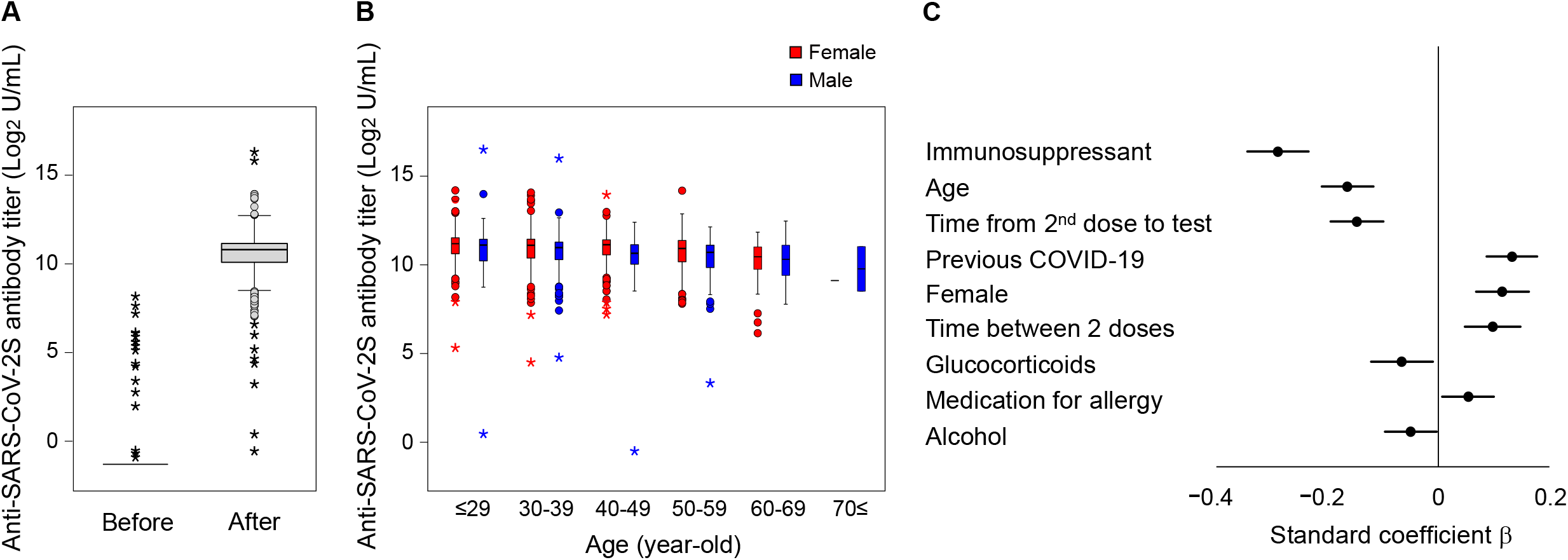
Distribution of antibody titer and multivariate linear regression model. **A, B.** Boxplots for the distribution of anti-SARS-CoV-2S antibody titers. A box represents interquartile range and a horizontal line in a box represents median. A vertical line represents a range excluding outliers. A circle and an asterisk represent outlier and extreme outlier, respectively. **A**. Before and after vaccination, in subjects whose data are available (n=1,774). Undetectable titers (<0.4 U/mL) are imputed with a value of 0.4 U/mL before log-transformed. **B**. After vaccination, according to age groups and sex. **C**. The variables retained in the final multivariate linear regression model to explain anti-SARS-CoV-2S antibody titers after vaccination. A dot and bar represents standardized coefficient β and 95% confidence interval for the variable.

We first performed univariate linear regression analyses to identify factors associated with the serum anti-SARS-CoV-2S antibody titer after vaccination. The factors significantly associated with a higher antibody titer were younger age (p<0.001), female (p<0.001), nurse (p<0.001), no previous/current smoking (p<0.001), previous COVID-19 (p<0.001), longer time between 2 doses (p<0.001), shorter time from 2^nd^ dose to sample collection (p<0.001), no immunosuppressive medication (p<0.001), no interstitial lung disease (p<0.001), no autoimmune disease (p<0.001), no hypertension (p=0.001), no medication for hypertension (p=0.001), exposure to COVID-19 patient (p=0.002), and less frequent alcohol consumption (p=0.012) (Supplementary Table S1).

We next performed a multivariate linear regression analysis using factors that showed a p-value <0.1 in univariate analyses. The factors retained in the final model (adjusted R^2^ 0.188) are immunosuppressive medication (p<0.001, standardized regression coefficient β −0.289 [95% confidence interval {CI} −0.344 – −0.234]), age (p<0.001, β −0.164 [95% CI −0.211 – −0.117]), time from 2^nd^ dose to sample collection (p<0.001, β −0.148 [95% CI − 0.195 – −0.100]), previous COVID-19 (p<0.001, β 0.133 [95% CI 0.087 – 0.178]), female (p<0.001, β 0.116 [95% CI 0.068 – 0.163]), time between 2 doses (p<0.001, β 0.100 [95% CI 0.050 – 0.150]), glucocorticoids (p=0.020, β −0.066 [95% CI −-0.121 – −0.010]), medication for allergy (p=0.024, β 0.054 [95% CI 0.007 – 0.100]), and alcohol (p=0.037, β −0.050 [95% CI −0.096 – −0.003]) (Figure 1C).

## Discussion

To our knowledge, this is the largest study to date to report the antibody responses to COVID-19 vaccination. In this study, all subjects who received 2 doses of BNT162b2 mRNA COVID-19 vaccine had a detectable level of serum anti-SARS-CoV-2S antibody, and all but one subject who were seronegative before vaccination became seropositive (99.9%). In addition, all of 18 subjects who were already seropositive before vaccination showed substantial antibody responses after the 2^nd^ dose. These results are consistent with previous smaller-scale studies [6, 7] and indicate that the vast majority of young-adult healthcare workers have good antibody responses following 2 doses of the BNT162b2 vaccine.

The large sample size of our study allowed for establishing a stable multivariate model to determine background factors that independently predict antibody responses. The strongest and the most significant factor was receiving immunosuppressive drugs (Figure 1C). Receiving glucocorticoids was also identified as an independent predictor (Figure 1C) even though our study target was mostly healthy workers and only 14 (0.9%) and 9 (0.6%) were taking glucocorticoids and immunosuppressant, respectively. Our data confirm the results of previous studies which demonstrated the reduced antibody responses among patients on immunosuppressive regimens [8].

Unexpectedly, medication for allergy was also identified as a factor significantly associated with higher antibody titers (Figure 1C). Although we have no information on the drug and diagnosis for the medication, we speculate that the majority were taking an anti-histamine drug for cedar pollen allergy, which is very common in Japan in spring. We also speculate that this may reflect some association between allergic diathesis and antibody responses. Some studies, on the other hand, have suggested possible therapeutic effects of histamine H1 receptor antagonists on COVID-19 [9]. Together with alcohol consumption as a negative predictor (Figure 1C), these novel associations deserve further investigation.

While only 10 participants (0.5%) in our study had a history of COVID-19, it was the fourth most significant factor in our multivariate model (Figure 1C). Its influence might have been underestimated since 2 participants who had the highest titers (93,200.0 and 65,800.0 U/mL) did not have a history of previous COVID-19 but both were seropositive before vaccination, and one had had close contact with an infected individual. Again, this result is consistent with previous reports [10, 11] and consolidates the evidence that the BNT162b2 vaccine induces more robust antibody responses in individuals previously infected with SARS-CoV-2.

Among demographic factors, age has been repeatedly reported to associate with reduced antibody responses after COVID-19 vaccination [6, 7]. Our study population is younger than those in previous studies and supplement the evidence. On the other hand, the sex difference has not been reported except for one study that showed a higher titer in women ≥80-year-old [7]. Our study with a larger sample size demonstrates for the first time significantly higher antibody titers in women than in men as a whole (median 2200.0 [IQR 1,350.0 – 2,807.5] vs. 1,805.0 U/mL [IQR 1,067.5 – 2,412.5], p<0.001).

Our data indicate that a longer interval between 2 doses within a range of 18-25 days is associated with a stronger antibody response. This may be interpreted as that the interval should not be shortened without justifiable reasons. Our data also show that a shorter period from the 2^nd^ dose to sample collection within a range of 14-32 days is associated with a stronger antibody response. This finding indicates that the peak of antibody responses takes place within 14 days of the 2^nd^ dose, which should be considered when interpreting the data.

Our study has some limitations. First, this is a single-center study in Japan with mostly Japanese subjects. Second, neutralizing antibody was not measured, although the assay we employed has been shown closely correlated with the titer of neutralizing antibody [12]. Third, most clinical information was collected with a questionnaire and cannot be verified. Nevertheless, we provide the largest data on antibody responses to the BNT162b2 mRNA COVID-19 vaccine in healthcare workers. Universally good responses demonstrated in our study further support the use of this vaccine in a wide range of populations and the predictive factors identified may help optimize the vaccination strategy and generate hypotheses for future studies.

## Data Availability

All data will be available after this manuscript is accepted by Scientific Journal.

## Acknowledgements

We thank all staff in Chiba University Hospital for supporting sample collection.

---

**Supplementary Table S1.** Background information and results of univariate linear regression analysis for post-vaccine antibody titer

| Variable | All<br>(n=2,015) |  | Post-vaccine antibody titer<br>available<br>(n=1,774) |  | Univariate linear regression<br>analysis for post-vaccine antibody<br>titer |  |
| --- | --- | --- | --- | --- | --- | --- |
| | Data available,<br>n | Value | Data available,<br>n | Value | Adjusted<br>coefficient $\beta$ | p-value |
| Age (year-old), median (IQR) | 2,015 | 37 (29-47) | 1,774 | 39.3 | -0.163 | <0.001 |
| Sex female, n (%) | 2,015 | 1,296 (64.3) | 1,774 | 1,168 (65.8) | -0.133 | <0.001 |
| Nationality Japanese, n (%) | 2,015 | 2,004 (99.5) | 1,774 | 1,765 (99.5) | 0.020 | 0.407 |
| Job category | 2,015 |  | 1,774 |  | 0.090* | <0.001 |
| Nurse |  | 672 (33.3) |  | 559 (31.5) |  |  |
| Doctor |  | 589 (29.2) |  | 494 (27.8) |  |  |
| Pharmacist |  | 58 (2.9) |  | 57 (3.2) |  |  |
| Dentist |  | 19 (0.9) |  | 11 (0.6) |  |  |
| Others |  | 677 (33.6) |  | 653 (36.8) |  |  |
| Body mass index category | 1,515 |  | 1,512 |  | 0.006 | 0.808 |
| <18.5 |  | 150 (9.9) |  | 150 (9.9) |  |  |
| 18.5-25 |  | 1,120 (73.9) |  | 1,118 (73.9) |  |  |
| $\geq 25$ | | 245 (16.2) | | 244 (16.1) | | |
| Smoking | 1,515 |  | 1,512 |  | -0.094 | <0.001 |
| Never, n (%) |  | 1,141 (75.3) |  | 1,139 (75.3) |  |  |
| Ex-smoker, n (%) |  | 323 (21.3) |  | 323 (21.4) |  |  |
| Current smoker, n (%) |  | 51 (3.4) |  | 50 (3.3) |  |  |
| Alcohol | 1,515 |  | 1,512 |  | -0.065 | 0.012 |
| No, n (%) |  | 417 (27.5) |  | 417 (27.6) |  |  |
| Sometimes, n (%) |  | 863 (57.0) |  | 860 (56.9) |  |  |
| Every day, n (%) |  | 235 (15.5) |  | 235 (15.5) |  |  |
| Comorbidities | 1,515 |  | 1,512 |  |  |  |
| Asthma, n (%) |  | 158 (10.4) |  | 158 (10.4) | -0.011 | 0.659 |
| Atopic dermatitis, n (%) |  | 134 (8.8) |  | 134 (8.9) | -0.002 | 0.944 |
| Hypertension, n (%) |  | 114 (7.5) |  | 114 (7.5) | -0.082 | 0.001 |
| Dyslipidemia, n (%) |  | 97 (6.4) |  | 97 (6.4) | -0.047 | 0.066 |
| Thyroid disease, n (%) |  | 55 (3.6) |  | 55 (3.6) | 0.016 | 0.546 |
| Malignancy, n (%) |  | 36 (2.4) |  | 36 (2.4) | −0.003 | 0.906 |
| Diabetes mellitus, n (%) |  | 25 (1.7) |  | 25 (1.7) | −0.045 | 0.082 |
| Autoimmune disease, n (%) |  | 13 (0.9) |  | 13 (0.9) | −0.218 | <0.001 |
| Ischemic heart disease, n (%) |  | 5 (0.3) |  | 5 (0.3) | −0.025 | 0.328 |
| Cerebral infarction, n (%) |  | 4 (0.3) |  | 4 (0.3) | 0.008 | 0.747 |
| Interstitial lung disease, n (%) |  | 2 (0.1) |  | 2 (0.1) | −0.177 | <0.001 |
| Chronic obstructive pulmonary disease, n (%) |  | 0 (0.0) |  | 0 (0.0) | NA | NA |
| Current medication | 1,515 |  | 1,512 |  |  |  |
| Allergy, n (%) |  | 188 (12.4) |  | 188 (12.4) | 0.045 | 0.083 |
| Hypertension, n (%) |  | 102 (6.7) |  | 102 (6.7) | −0.090 | <0.001 |
| Dyslipidemia, n (%) |  | 77 (5.1) |  | 77 (5.1) | −0.023 | 0.366 |
| Inhaled corticosteroid, n (%) |  | 32 (2.1) |  | 32 (2.1) | −0.039 | 0.130 |
| Thyroid disease, n (%) |  | 27 (1.8) |  | 27 (1.8) | −0.003 | 0.901 |
| Diabetes mellitus, n (%) |  | 20 (1.3) |  | 20 (1.3) | −0.019 | 0.471 |
| Glucocorticoids, n (%) |  | 14 (0.9) |  | 14 (0.9) | −0.207 | <0.001 |
| Immunosuppressant, n (%) |  | 9 (0.6) |  | 9 (0.6) | −0.299 | <0.001 |
| Insulin, n (%) |  | 3 (0.2) |  | 3 (0.2) | −0.038 | 0.140 |
| Antimicrobial, n (%) |  | 3 (0.2) |  | 3 (0.2) | −0.043 | 0.096 |
| Previous COVID-19, n (%) | 2,015 | 10 (0.5) | 1,774 | 9 (0.5) | 0.115 | <0.001 |
| Flu symptoms within a year, n (%) | 1,515 | 539 (35.6) | 1,512 | 539 (35.6) | 0.012 | 0.650 |
| Exposure to COVID-19 patient | 1,515 |  | 1,512 |  | 0.078 | 0.002 |
| Hardly, n (%) |  | 1,333 (88.0) |  | 1,331 (88.0) |  |  |
| < 15 minutes, n (%) |  | 76 (5.0) |  | 76 (5.0) |  |  |
| ≥ 15 minutes, n (%) |  | 106 (7.0) |  | 105 (6.9) |  |  |
| Time between 1<sup>st</sup> and 2<sup>nd</sup> doses (day), mean (SD) | 1,987 | 21.2 (0.7) | 1,774 | 21.1 (0.6) | 0.106 | <0.001 |
| Time between 2<sup>nd</sup> dose and sample collection (day), median (IQR) | 1,774 | 15 (14-21) | 1,774 | 15 (14-21) | −0.161 | <0.001 |

\*Nurse vs. non-nurse. Other combinations did not yield p-value<0.1. IQR, interquartile range; SD, standard deviation.

